# A retrospective study evaluating efficacy and safety of compassionate use of tocilizumab in 13 patients with severe-to-critically ill COVID-19: analysis of well-responding cases and rapidly-worsening cases after tocilizumab administration

**DOI:** 10.1101/2020.06.24.20134288

**Authors:** Shoji Hashimoto, Heita Kitajima, Tsuyoshi Arai, Yoshitaka Tamura, Takayuki Nagai, Hiroshi Morishita, Hiroto Matsuoka, Yuki Han, Seijiro Minamoto, Tomonori Hirashima, Tomoki Yamada, Yozo Kashiwa, Makoto Kameda, Seiji Yamaguchi, Kazuko Uno, Emi Nakayama, Tatsuo Shioda, Kazuyuki Yoshizaki, Sujin Kang, Tadamitsu Kishimoto, Toshio Tanaka

## Abstract

We administered tocilizumab into 13 severe-to-critically ill patients with coronavirus disease 2019 (COVID-19) for compassionate use in combination with potential anti-viral agents in those who required an oxygen supply and showed increased laboratory inflammatory markers such as C-reactive protein (CRP) and ferritin. One injection of tocilizumab led to rapid improvements in clinical features, inflammatory findings, and oxygen supply in seven patients with severe COVID-19 and substantial amelioration in two patients who were critically ill, whereas four patients, who exhibited rapidly worsened respiratory function, required artificial ventilatory support even after tocilizumab treatment. Three of these four patients ultimately recovered from deterioration after methylprednisolone treatment. Administration of tocilizumab did not affect viral elimination nor IgG production specific for the virus. Compared with well-responding patients, rapidly-worsened patients showed a significantly higher ratio of ferritin vs. CRP. These findings suggest that tocilizumab has beneficial effects in severe-to-critically ill patients with COVID-19; however, in some cases, addition of methylprednisolone is required for disease rescue.

## Introduction

Coronavirus disease 2019 (COVID-19), caused by severe acute respiratory syndromes coronavirus-2 (SARS-CoV-2), has rapidly spread worldwide.^1-4^ By the end of May 2020, more than 6 million people were diagnosed with COVID-19, with a mortality rate of approximately 6%. Thus, vaccines and therapeutic drugs are urgently needed to stop the spread of the disease and decrease mortality; however, no vaccines and drugs except for remdesivir as an emergency use drug have been developed and approved.^5^ Various studies have indicated that a cytokine storm, also known as hyperinflammation, is a pathological mechanism underlying the development of severe disease, leading to a critically ill state of patients including acute respiratory distress syndrome, multiple organ dysfunction syndrome, and shock.^6-8^ Among the cytokines involved in severe and critical cases of COVID-19, interleukin (IL)-6 is highly elevated and can be used a prognostic marker.^9-11^ Additionally, IL-6 plays a major pathological role in disease worsening. Several case series and case reports have demonstrated the beneficial effects of tocilizumab, a humanized anti-IL-6 receptor monoclonal antibody, in patients with COVID-19.^12-16^ Based on recent findings regarding the clinical and laboratory features of COVID-19, ^1-4,9,10^ our hospital developed a therapeutic protocol for managing COVID-19 and used tocilizumab to treat severe-to-critically ill patients with COVID-19. Here, we report our experience using tocilizumab to treat patients with COVID-19.

## Patients and methods

### Patients with COVID-19

Seventy patients with COVID-19 were admitted to our hospital by the end of May 2020. Among them, 13 patients were diagnosed as severe-to-critically ill, as they required oxygen supply because of severe pneumonia and were intravenously administered tocilizumab at 400 mg once in combination with anti-SARS-CoV-2 drugs such as lopinavir/ritonavir, ciclesonide, or favipiravir. All patients provided written informed consent, and the off-label compassionate use of tocilizumab was approved by the Ethics Committee of Osaka Habikino Medical Center (Approved ID: 150-7).

### Protocol of the indication of tocilizumab for patients with COVID-19

Based on various previous reports of characterizations patients with COVID-19, the inclusion criteria of off-label indication of tocilizumab were set as follows:

1. Elevated inflammatory findings: C-reactive protein (CRP) level >5 mg/dL or ferritin >1000 ng/mL
2. Requirement of oxygen supply or rapid progression according to chest image evaluation (more than 50% increase in infiltrates over 24-48 hours)

Patients with elevated procalcitonin and patients who also had bacterial infections were excluded.

### Quantification of viral RNA

Viral RNA was isolated from 70 *µ*L of serum using a QIAmp Viral RNA Mini kit (Qiagen), according to the manufacturers’ protocol. Viral RNA was quantified using a One-StepPrimeScript III RT-PCR kit (TaKaRa), and the following universal primers for N2 region of SARS-CoV-2: NIID_2019-nCOV_N_F2 AAATTTTGGGGACCAGGAAC and NIID_2019-nCOV_N_R2 TGGCAGCTGTGTAGGTCAAC with NIID_2019-nCOV_N_P2 probe: FAM-ATGTCGCGCATTGGCATGGA-TAMRA. Five *µ*L of extracted RNA was used for the reaction. The PCR conditions were 25°C for 10 min for activation, 52°C for 5 min for reverse transcription, 95°C 10 sec for inactivation, followed by 45 cycles of 95°C for 5 s, and 20°C for 30 s. The fluorescent signals were detected with a QuantiStudio 3 Real-Time PCR System (Applied Biosystems).

### Detection of specific antibody against SARS-CoV-2

The serum levels of IgM and IgG class antibodies specific for SARS-CoV-2 were determined by using the Corona Virus COVID-19 Antibody Rapid Detection kit according to the manufacturer’s instructions (Healgen Scientific Ltd.).^17^ The test samples in an individual patient included all of the sera collected before and 1-2 and 3-4 weeks after tocilizumab injection, or in some cases at 4-5 weeks after injection, depending on the availability.

### Statistical analysis

The significance of the difference between well-responding group and rapidly-worsening group was evaluated using the Man-Whitney U test. A value of *P* <0.05 was considered as statistically significant.

## Results

### Clinical outcome of tocilizumab administration

The patients’ characteristic features and clinical courses are shown in **Table 1**. Eleven male patients and two female patients with a mean age of 63 years were evaluated. Five patients were complicated with diabetes mellitus, five with hypertension, and two with chronic obstructive pulmonary disease. At admission, two patients were critically ill and required artificial ventilator management before tocilizumab injection, and severe disease was diagnosed in 11 patients. The clinical course of each patient is shown in **Table 1**. Tocilizumab caused no adverse events. Seven patients promptly recovered from fever and malaise and lowered their oxygen support, and were free of oxygen support within a week on average (well-responding group). PCR analysis of the nasopharyngeal specimens showed negative results for SARS-CoV-2 10-25 days (15.4 days as average) and the patients were discharged 12-27 days (17 days as average) after tocilizumab administration. However, four patients showed further worsening of respiratory function and required artificial ventilatory support (rapidly-worsening group); these patients were administered methylprednisolone and recovered from such a support within a week. Two patients were discharged from the hospital, and one patient died because of sudden laryngotracheal stenosis. Two patients who were critically ill and required artificial ventilator management before tocilizumab injection were ameliorated in respiratory function. Analysis of clinical outcomes showed that by 1 week after tocilizumab treatment, eight (62%), 1 (8%), and 4 (31%) patients improved, had no change, and worsened, respectively, whereas nine (69%), 3 (23%), and 1 patient (8%) were cured, improved, and died, respectively, by 1 month after treatment. A representative case that responded well to tocilizumab and a case that worsened despite tocilizumab injection are shown in **Figures 1** and **2**, respectively. The changes in laboratory parameters are shown in **Figure 3**. Tocilizumab injection rapidly decreased serum CRP levels followed by a gradual decrease in ferritin and increase in the number of peripheral lymphocytes.

**Table 1.**
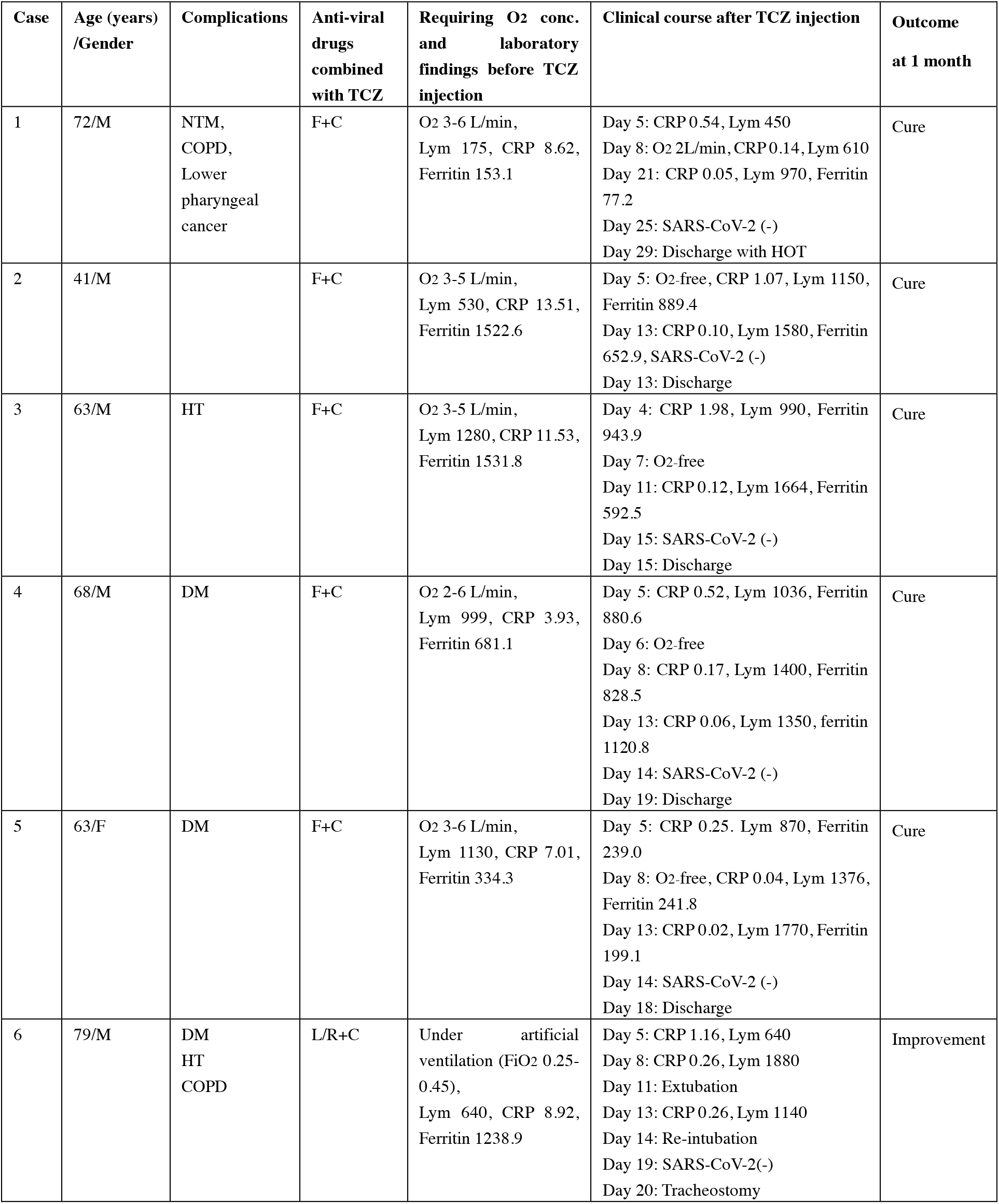
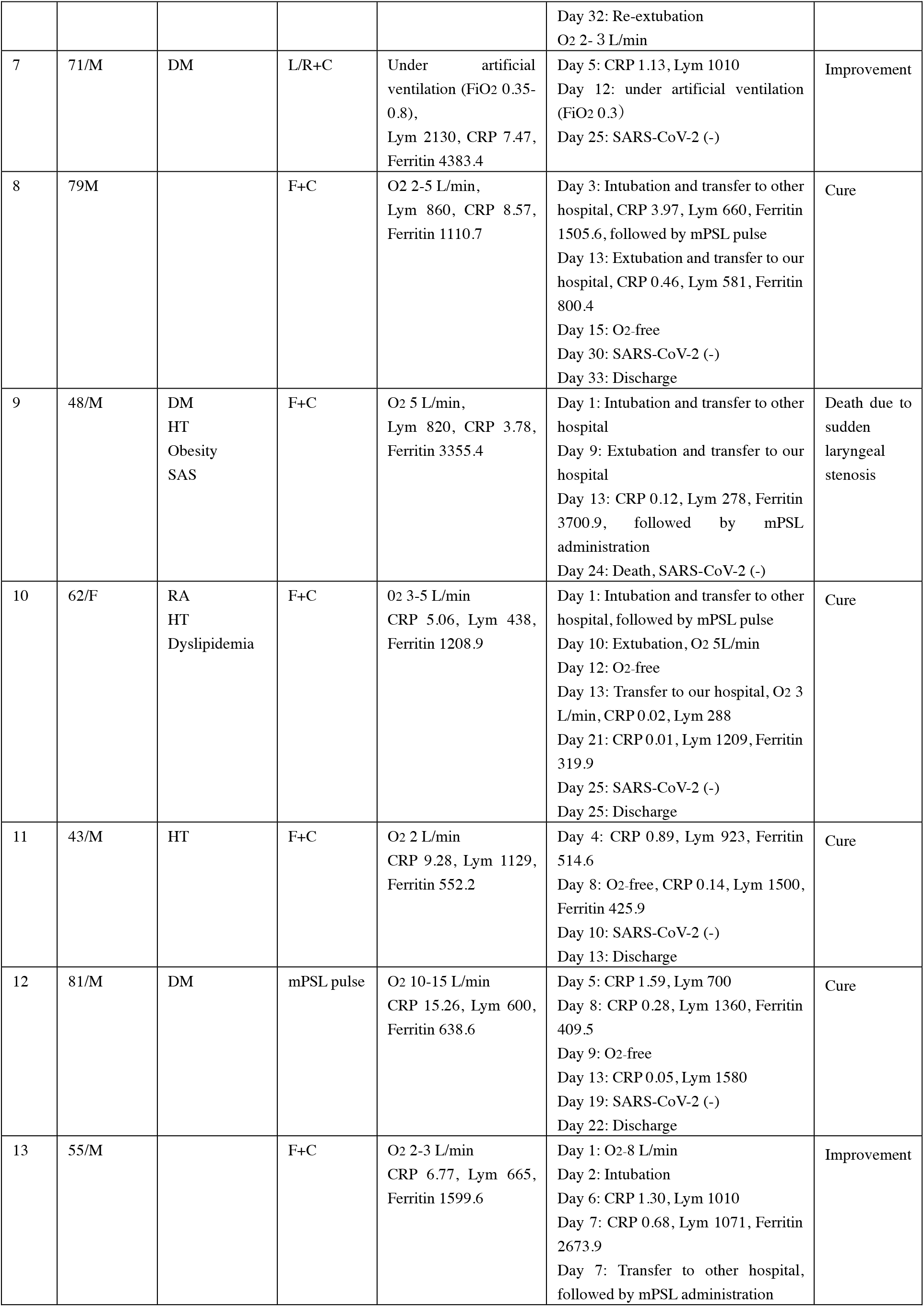
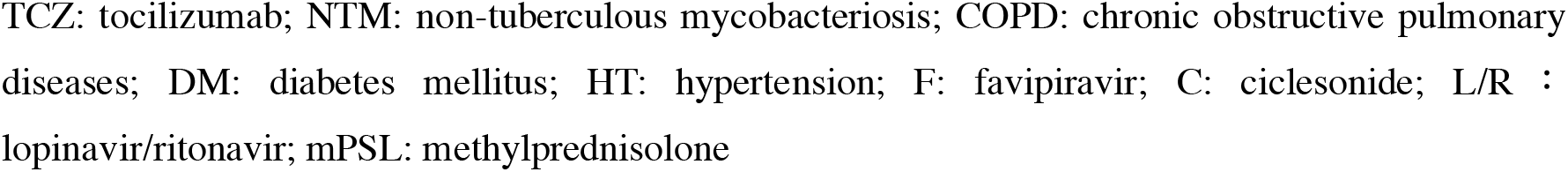
Characteristic features and clinical course of patients with COVID-19 treated with tocilizumab.

| Case | Age (years) /Gender | Complications | Anti-viral drugs combined with TCZ | Requiring O<sub>2</sub> conc. and laboratory findings before TCZ injection | Clinical course after TCZ injection | Outcome at 1 month |
| --- | --- | --- | --- | --- | --- | --- |
| 1 | 72/M | NTM, COPD, Lower pharyngeal cancer | F+C | O<sub>2</sub> 3-6 L/min, Lym 175, CRP 8.62, Ferritin 153.1 | Day 5: CRP 0.54, Lym 450<br>Day 8: O<sub>2</sub> 2L/min, CRP 0.14, Lym 610<br>Day 21: CRP 0.05, Lym 970, Ferritin 77.2<br>Day 25: SARS-CoV-2 (-)<br>Day 29: Discharge with HOT | Cure |
| 2 | 41/M |  | F+C | O<sub>2</sub> 3-5 L/min, Lym 530, CRP 13.51, Ferritin 1522.6 | Day 5: O<sub>2</sub>-free, CRP 1.07, Lym 1150, Ferritin 889.4<br>Day 13: CRP 0.10, Lym 1580, Ferritin 652.9, SARS-CoV-2 (-)<br>Day 13: Discharge | Cure |
| 3 | 63/M | HT | F+C | O<sub>2</sub> 3-5 L/min, Lym 1280, CRP 11.53, Ferritin 1531.8 | Day 4: CRP 1.98, Lym 990, Ferritin 943.9<br>Day 7: O<sub>2</sub>-free<br>Day 11: CRP 0.12, Lym 1664, Ferritin 592.5<br>Day 15: SARS-CoV-2 (-)<br>Day 15: Discharge | Cure |
| 4 | 68/M | DM | F+C | O<sub>2</sub> 2-6 L/min, Lym 999, CRP 3.93, Ferritin 681.1 | Day 5: CRP 0.52, Lym 1036, Ferritin 880.6<br>Day 6: O<sub>2</sub>-free<br>Day 8: CRP 0.17, Lym 1400, Ferritin 828.5<br>Day 13: CRP 0.06, Lym 1350, ferritin 1120.8<br>Day 14: SARS-CoV-2 (-)<br>Day 19: Discharge | Cure |
| 5 | 63/F | DM | F+C | O<sub>2</sub> 3-6 L/min, Lym 1130, CRP 7.01, Ferritin 334.3 | Day 5: CRP 0.25, Lym 870, Ferritin 239.0<br>Day 8: O<sub>2</sub>-free, CRP 0.04, Lym 1376, Ferritin 241.8<br>Day 13: CRP 0.02, Lym 1770, Ferritin 199.1<br>Day 14: SARS-CoV-2 (-)<br>Day 18: Discharge | Cure |
| 6 | 79/M | DM<br>HT<br>COPD | L/R+C | Under artificial ventilation (FiO<sub>2</sub> 0.25-0.45), Lym 640, CRP 8.92, Ferritin 1238.9 | Day 5: CRP 1.16, Lym 640<br>Day 8: CRP 0.26, Lym 1880<br>Day 11: Extubation<br>Day 13: CRP 0.26, Lym 1140<br>Day 14: Re-intubation<br>Day 19: SARS-CoV-2(-)<br>Day 20: Tracheostomy | Improvement |
|  |  |  |  |  | Day 32: Re-extubation<br>O2 2- 3 L/min |  |
| 7 | 71/M | DM | L/R+C | Under artificial ventilation (FiO2 0.35-0.8),<br>Lym 2130, CRP 7.47, Ferritin 4383.4 | Day 5: CRP 1.13, Lym 1010<br>Day 12: under artificial ventilation (FiO2 0.3)<br>Day 25: SARS-CoV-2 (-) | Improvement |
| 8 | 79M |  | F+C | O2 2-5 L/min,<br>Lym 860, CRP 8.57, Ferritin 1110.7 | Day 3: Intubation and transfer to other hospital, CRP 3.97, Lym 660, Ferritin 1505.6, followed by mPSL pulse<br>Day 13: Extubation and transfer to our hospital, CRP 0.46, Lym 581, Ferritin 800.4<br>Day 15: O2-free<br>Day 30: SARS-CoV-2 (-)<br>Day 33: Discharge | Cure |
| 9 | 48/M | DM<br>HT<br>Obesity<br>SAS | F+C | O2 5 L/min,<br>Lym 820, CRP 3.78, Ferritin 3355.4 | Day 1: Intubation and transfer to other hospital<br>Day 9: Extubation and transfer to our hospital<br>Day 13: CRP 0.12, Lym 278, Ferritin 3700.9, followed by mPSL administration<br>Day 24: Death, SARS-CoV-2 (-) | Death due to sudden laryngeal stenosis |
| 10 | 62/F | RA<br>HT<br>Dyslipidemia | F+C | O2 3-5 L/min<br>CRP 5.06, Lym 438, Ferritin 1208.9 | Day 1: Intubation and transfer to other hospital, followed by mPSL pulse<br>Day 10: Extubation, O2 5L/min<br>Day 12: O2-free<br>Day 13: Transfer to our hospital, O2 3 L/min, CRP 0.02, Lym 288<br>Day 21: CRP 0.01, Lym 1209, Ferritin 319.9<br>Day 25: SARS-CoV-2 (-)<br>Day 25: Discharge | Cure |
| 11 | 43/M | HT | F+C | O2 2 L/min<br>CRP 9.28, Lym 1129, Ferritin 552.2 | Day 4: CRP 0.89, Lym 923, Ferritin 514.6<br>Day 8: O2-free, CRP 0.14, Lym 1500, Ferritin 425.9<br>Day 10: SARS-CoV-2 (-)<br>Day 13: Discharge | Cure |
| 12 | 81/M | DM | mPSL pulse | O2 10-15 L/min<br>CRP 15.26, Lym 600, Ferritin 638.6 | Day 5: CRP 1.59, Lym 700<br>Day 8: CRP 0.28, Lym 1360, Ferritin 409.5<br>Day 9: O2-free<br>Day 13: CRP 0.05, Lym 1580<br>Day 19: SARS-CoV-2 (-)<br>Day 22: Discharge | Cure |
| 13 | 55/M |  | F+C | O2 2-3 L/min<br>CRP 6.77, Lym 665, Ferritin 1599.6 | Day 1: O2-8 L/min<br>Day 2: Intubation<br>Day 6: CRP 1.30, Lym 1010<br>Day 7: CRP 0.68, Lym 1071, Ferritin 2673.9<br>Day 7: Transfer to other hospital, followed by mPSL administration | Improvement |

TCZ: tocilizumab; NTM: non-tuberculous mycobacteriosis; COPD: chronic obstructive pulmonary diseases; DM: diabetes mellitus; HT: hypertension; F: favipiravir; C: ciclesonide; L/R : lopinavir/ritonavir; mPSL: methylprednisolone

**Figure 1.**
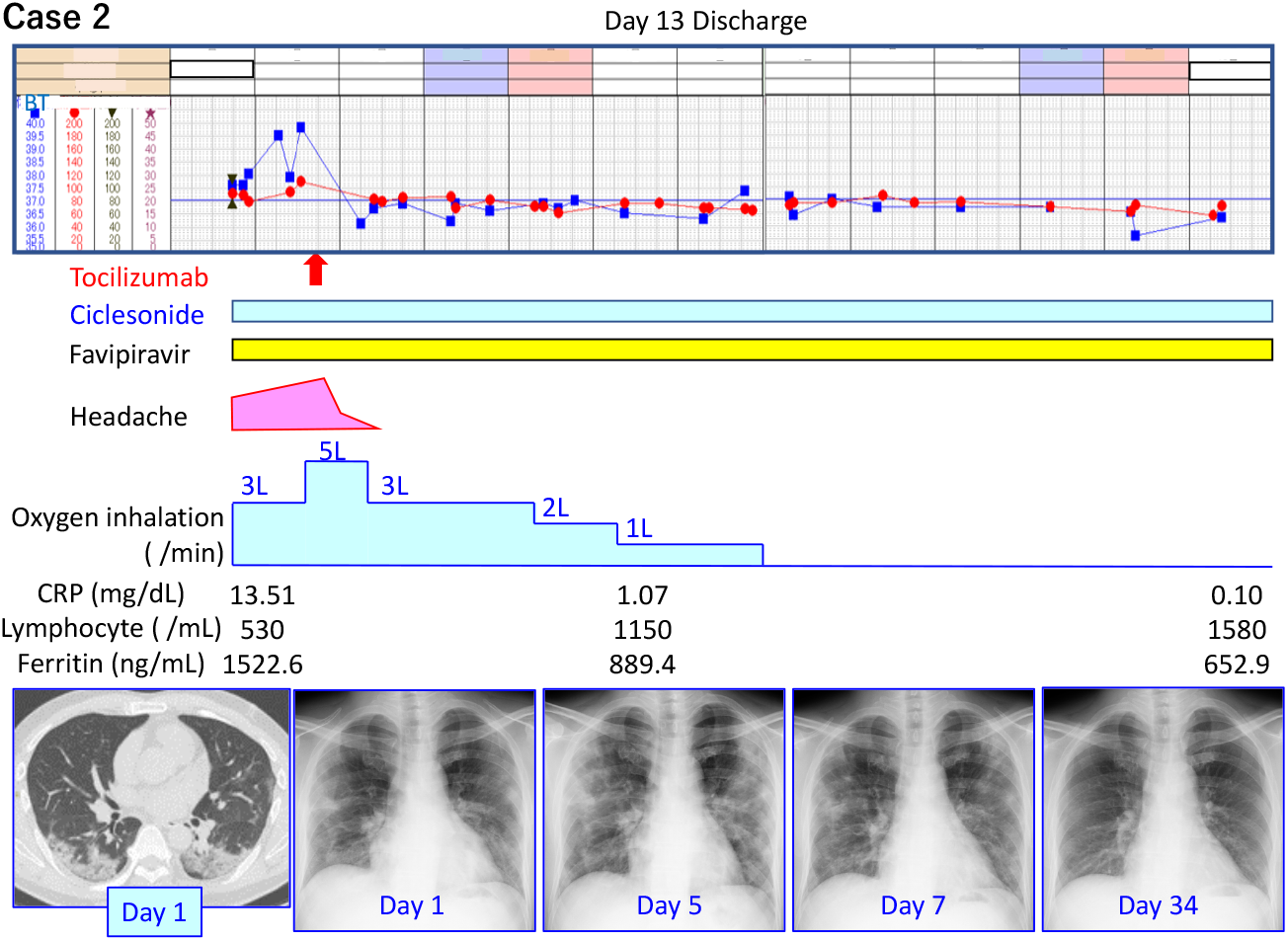
One representative case that responded well to tocilizumab administration.

**Figure 2.**
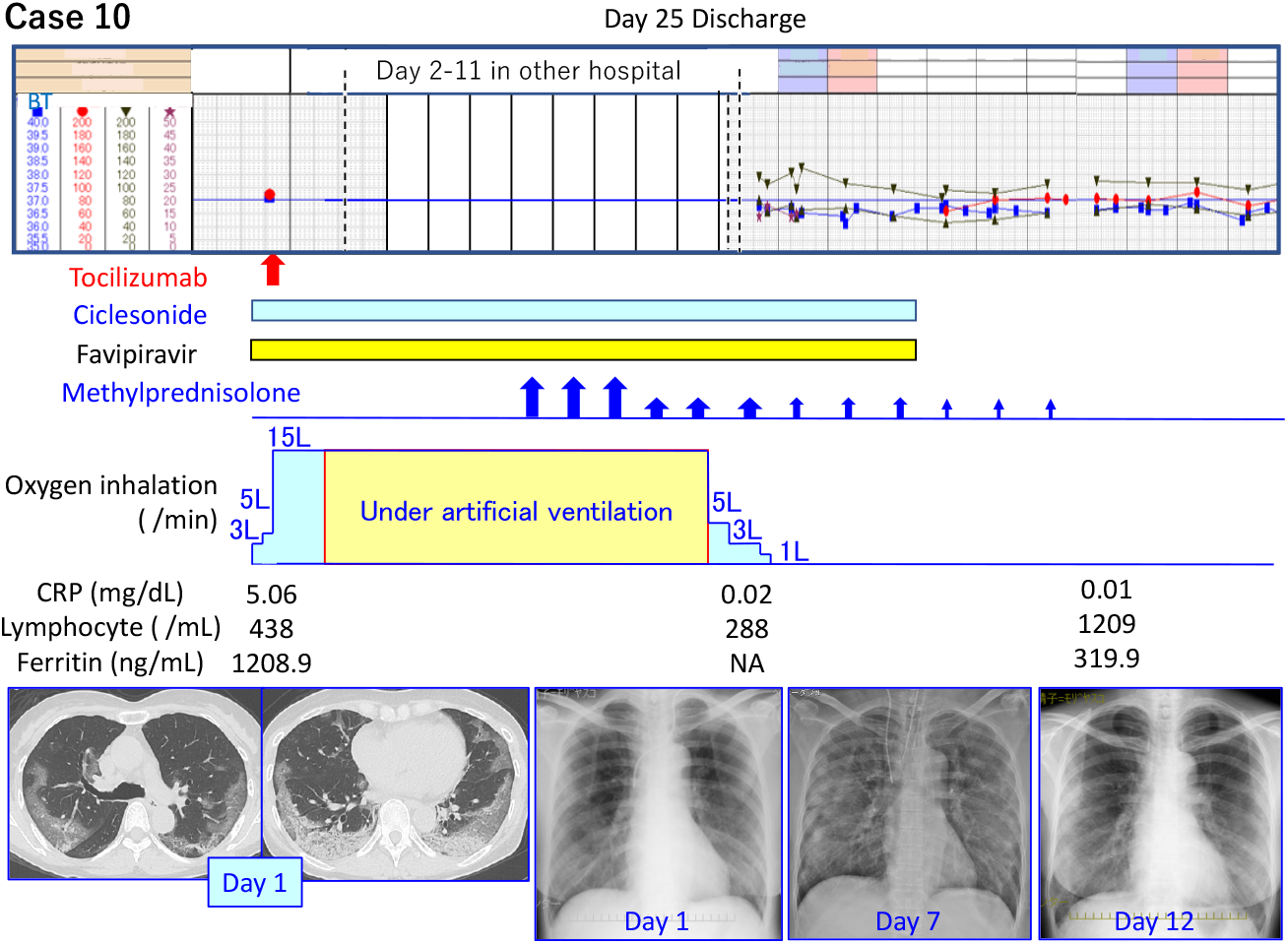
One representative case that worsened rapidly after tocilizumab administration.

**Figure 3.**
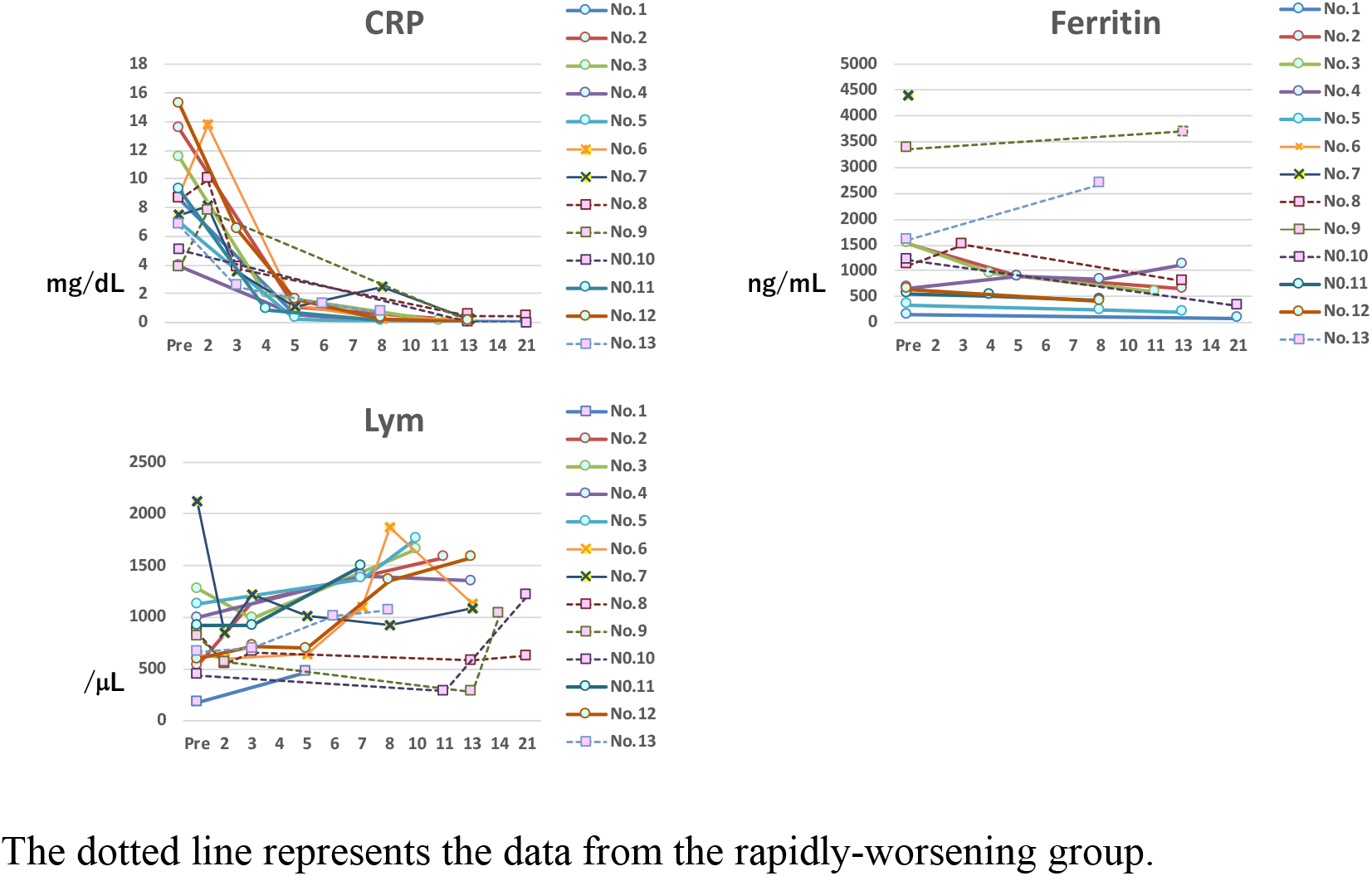
Changes in CRP, ferritin, and the number of peripheral lymphocytes before and after tocilizumab injection.

### Difference between well-responding group and rapidly-worsening group

Respiratory disturbance acutely progressed in some patients with severe disease.^1-3^ The clear difference in the clinical features between the well-responding group and the rapidly-worsening group was not observed. In the laboratory results, there was no difference in the basal levels of CRP and ferritin or in number of peripheral lymphocytes between groups, whereas the ratio of ferritin/CRP was significantly higher in the rapidly-worsening group than in the well-responding group (373.1±346.8 vs. 83.7±56.7, *p* = 0.01) (**Table 2**).

**Table 2.**
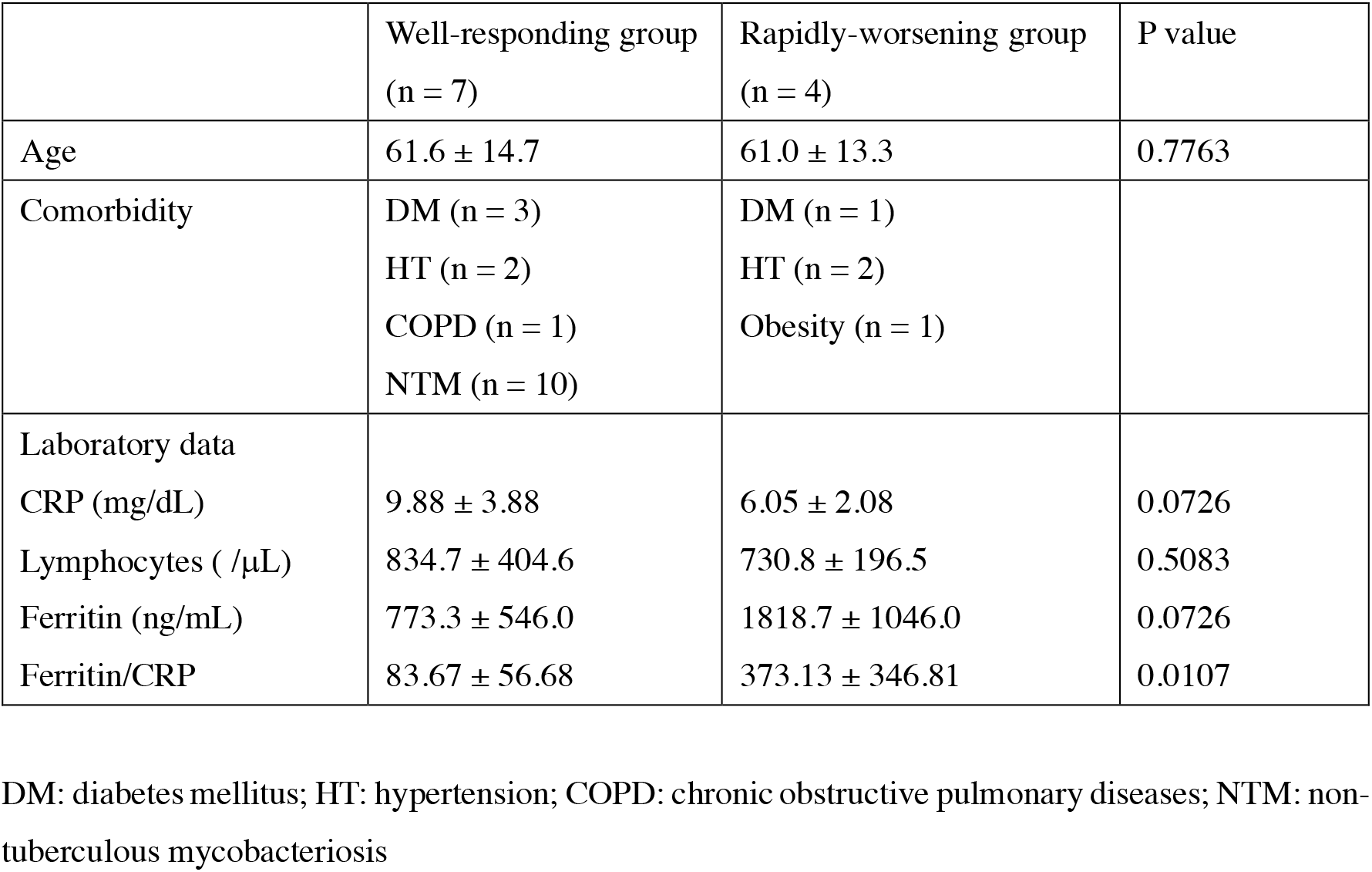
Comparative features before tocilizumab administration between well- responding group and rapidly-worsening group.

### Effect of tocilizumab injection on viral elimination and antibody production

Because IL-6 plays a crucial role in the host defense against pathogens and promotes T cell and B cell activation and differentiation,^18,19^ we next evaluated whether tocilizumab injection affects elimination of SARS-CoV-2 and specific antibody production. Three out of nine patients tested showed viremia, and their viral load transiently increased at 2-3 days after tocilizumab injection but then decreased (**Figure 4**). In addition, we detected IgG class antibody in all nine patients, whose sera were available for the test at 1-2 weeks after tocilizumab administration. These results suggest that tocilizumab in combination with anti-viral drugs did not suppress viral elimination or induction of the IgG class antibody specific for SARS-CoV-2.

**Figure 4.**
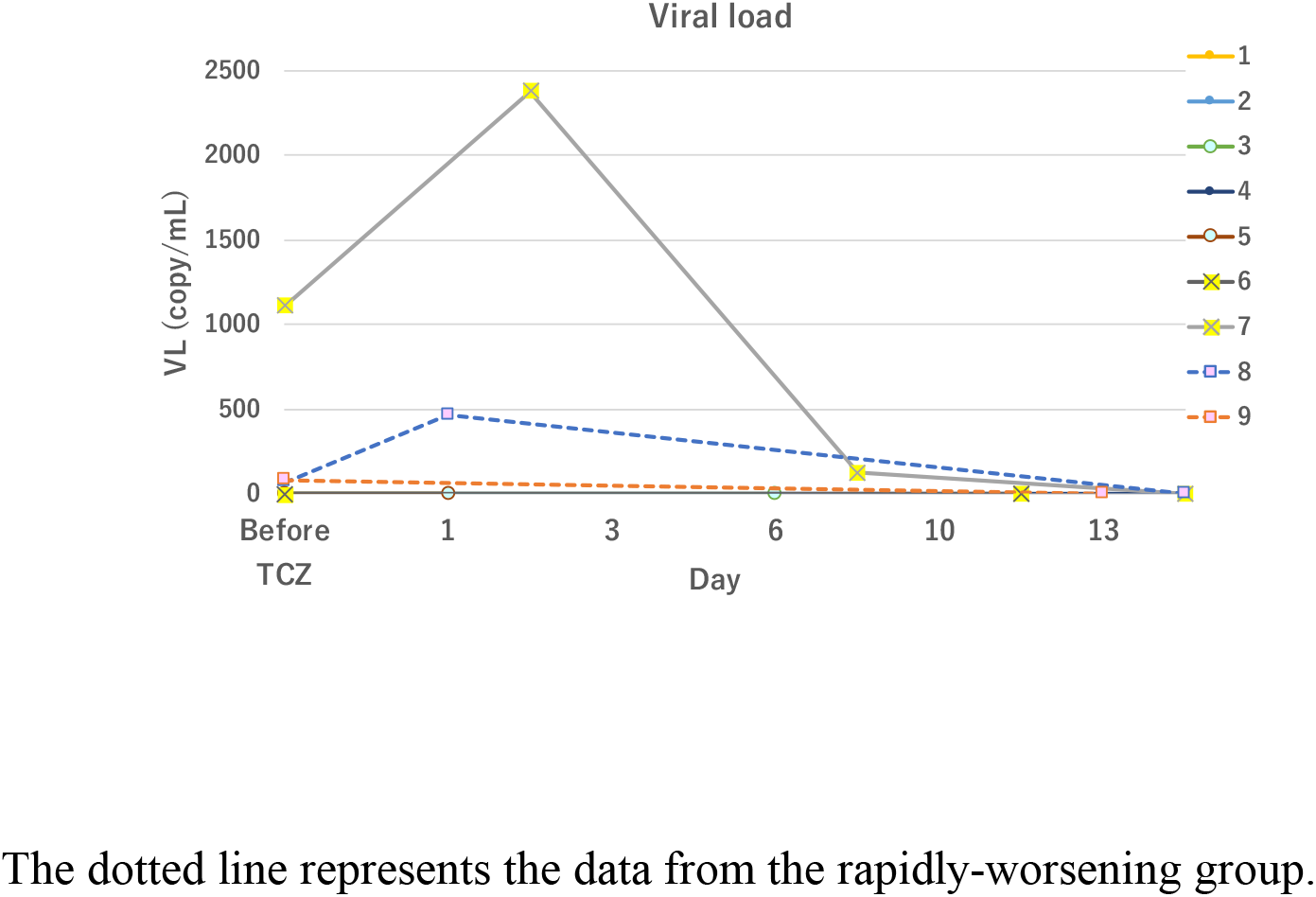
Changes in SARS-CoV-2 load before and after tocilizumab injection.

## Discussion

We describe our experience in which tocilizumab was administered to 13 severe-to- critically ill patients with COVID-19. Seven patients promptly improved in response to tocilizumab (well-responding group), whereas four patients showed worsened respiratory function and required artificial ventilation (rapidly-worsening group). Recent various case series and case reports have demonstrated the promising efficacy of tocilizumab for severe-to-critically ill COVID-19 either monotherapy or its combination with corticosteroids,^12-16,20-23^ although opposite results were observed in some other studies.^24,25^ According to our experience with 13 patients with COVID-19 patients, it is due to the clinical status and the timing of tocilizumab administration.

Compared with patients with non-severe COVID-19, those with severe-to-critically ill COVID-19 showed lymphocytopenia, elevated CRP, ferritin and D-dimer.^6,7,11,26^ Patients in the rapidly-worsening group showed a significantly higher ratio of ferritin/CRP, perhaps reflecting a more severe cytokine storm. CRP is predominantly regulated by IL-6; tocilizumab, as previously demonstrated in rheumatoid arthritis and Castleman disease,^27^ caused a prompt reduction in CRP levels in all patients with COVID-19 and a gradual reduction in ferritin levels along with an increase in the number of peripheral lymphocytes. Ferritin production is regulated by proinflammatory cytokines and IL-18,^28^ and thus high elevation of ferritin indicates exaggerated production of other cytokines in addition to IL-6. Similar effects occur in macrophage activation syndrome (MAS) complicated with systemic juvenile idiopathic arthritis (sJIA) and secondary haemophagocytic lymphohistiocytosis,^6,18,29^ in which ferritin is highly elevated, associated with the storm of various cytokines, including IL-1, IL-2, IL-6, IL-18, and IFN-γ, and others. Although the incidence of MAS in patients with sJIA appears to be lower during tocilizumab treatment, tocilizumab does not completely suppress the onset of the condition. In such cases, prednisolone and cyclosporine are generally used. Similar to this situation, four patients in the rapidly-worsening group were administered methylprednisolone and recovered from artificial ventilatory support. Thus, as immunosuppressive therapeutic treatments, tocilizumab monotherapy or its combination with methylprednisolone may depend on the patient’s immunological status, and the ratio of ferritin/CRP may be a useful marker for selecting the treatment procedure, although additional studies are essential to confirm this.

IL-6 is a cytokine that helps maintain homeostasis. When infections occur, IL-6 is promptly produced and plays a major role against infectious agents by producing acute phase proteins and activating T cells and B cells, leading to their differentiation into effector T cells and Ig production, respectively.^18,19^ Therefore, an important concern is that tocilizumab may suppress viral elimination and humoral immunity for the virus. However, our experience suggests that this is not the case when tocilizumab is administered with potential anti-viral drugs. Rather, flow cytometric analysis of immune cells from patients with COVID-19 showed that impaired immune cell cytotoxicity in severe cases was IL-6-dependent and thus targeting IL-6 may restore anti-viral activity.^30^

This report has several limitations. The sample size was small, and the data were analyzed retrospectively. Moreover, the treatment protocol involved combination therapy of tocilizumab with anti-viral drugs, so the direct effect of tocilizumab could not be evaluated. Further evaluation using a randomized controlled trial design is essential.

## Data Availability

The availability of all data were included in the manuscript.

## Reference

1. Huang C, Wang Y, Li X, Ren L, Zhao J, Hu Y, et al. Clinical features of patients infected with 2019 novel coronavirus in Wuhan, China. Lancet 2020 Feb 15:395(10223):497–506.

2. Wang D, Hu B, Hu C, Zhu F, Liu X, Zhang J, et al. Clinical characteristics of 138 hospitalized patients with 2019 novel coronavirus-infected pneumonia in Wuhan, China. JAMA 2020 Feb 7. doi:10.1001/jama.2020.1585. [Epub ahead of print]

3. Wu Z, McGoogan JM. Characteristics of and important lessons from the coronavirus disease 2019 (COVID-2019) outbreak in China: Summary of a report of 72314 cases from the Chinese Center for Disease Control and Prevention. JAMA 2020 Feb 24. doi:10.1001/jama.2020.2648. [Epub ahead of print]

4. World Health Organization. Coronavirus disease 2019 (COVID-19) situation report-51. Geneva, Switzerland: World Health Organization; 2020. http://www.who.int/docs/default-source/coronaviruse/situation-reports/20200311-sitrep-51-covid-19.

5. McCreary EK, Pogue JM. COVID-19 treatment: a review of early and emerging options. Open Forum Infectious Diseases 2020;7(4), ofaa105. https://doi.org/10.1093/ofid/ofaa105.

6. Mehta P, McAuley DF, Brown M, Sanchez E, Tattersall RS, Manson JJ; HLH across specialist collaboration, UK. COVID-19: consider cytokine storm syndromes and immunosuppression. Lancet 2020;395(10299):1033–1034.

7. Ye Q, Wang B, Mao J. The pathogenesis and treatment of the ‘Cytokine Storm’ in COVID-19. J Infect. 2020;80:607–613.

8. Moore JB, June CH. Cytokine release syndrome in severe COVID-19. Science 10.1126/science.abb8925(2020).

9. Zhou F, Yu T, Du R, Fan G, Liu Y, Liu Z, et al. Clinical course and risk factors for mortality of adult inpatients with COVID-19 in Wuhan, China: a retrospective cohort study. Lancet. 2020;395(10229):1054–1062.

10. Yang X, Yu Y, Xu J, Shu H, Xia J, Liu H, et al. Clinical course and outcomes of critically ill patients with SARS-CoV-2 pneumonia in Wuhan, China: a single-centered retrospective, observational study. Lancet Respir Med. 2020;8(5):475–481.

11. Liu T, Zhang J, Yang Y, Ma H, Li Z, Zhang J, et al. The role of interleukin-6 in monitoring severe case of coronavirus disease 2019. EMBO Mol Med. 2020 May 19. doi: 10.15252/emmm.202012421.

12. Xu X, Han M, Li T, Sun W, Wang D, Fu B, Zhou Y. et al. Effective treatment of severe COVID-19 patients with tocilizumab. PNAS. 2000;117(20):10970–10975.

13. Luo P, Liu Y, Qia L, Liu X, Liu D, Li J. Tocilizumab treatment in COVID-19: A single center experience. J Med Virol. 2020 April 6:10.1002/jmv.25801. doi: 10.1002/jmv.25801.

14. Michot J, Albiges L, Chaput N, Saada V, Pommeret F, Griscelli F, et al. Tocilizumab, an anti-IL-6 receptor antibody, to treat Covid-19-related respiratory failure: A case report. Ann Oncol. 2020 Apr 2;S0923-7534(20)36387-0. Doi: 10.1016/j.annonc.2020.03.300.

15. Zhang X, Song K, Tong F, Fei M, Guo H, Lu Z, et al. First case of COVID-19 in a patient with multiple myeloma successfully treated with tocilizumab. Blood Adv. 2020;4(7):1307–1310.

16. Di Giambenedetto S, Ciccullo A, Borghetti A, Gambassi G, Landi F, Visconti E, et al. Off-label use of tocilizumab in patients with SARS-CoV-2 infection. J Med Virol. 2020 April 16;10.1002/jmv.25897. doi: 10.1002/jmv.25897.

17. https://www.healgen.com/if-respiratory-covid-19

18. Tanaka T, Narazaki M, Kishimoto T. Immunotherapeutic implications of IL-6 blockade for cytokine storm. Immunotherapy 2016;8:959–970.

19. Kang S, Tanaka T, Narazaki M, Kishimoto T. Targeting interleukin-6 signaling in clinic. Immunity 2019;50:1007–1023.

20. Sciascia S, Apra F, Baffa A, Baldovino S, Boaro D, Boero R, et al. Pilot prospective open, single-arm multicenter study on off-label use of tocilizumab in severe patients with COVID-19. Clin Exp Rheumatol. 2020;38(3):529–532.

21. Alattar R, Ibrahim TBH, Shaar SH, Abdalla S, Shukri K, Daghfai JD, et al. Tocilizumab for the treatment of severe coronavirus diseases 2019. J Med Virol. 2020 May 5;10.1002/jmv.25964. doi: 10.1002/jmv.25964.

22. Toniati P, Piva S, Cattallni M, Garrata E, Regola F, Castelli F, et al. Tocilizumab for the treatment of severe COVID-19 pneumonia with hyperinflammatory syndrome and acute respiratory failure: a single center study of 100 patients in Brescia, Italy. Autoimmun Rev. 2020;19(7):102568.

23. Klopfenstein T, Zayet S, Lohse A, Balblanc J-C, Badie J, Royer P-Y., et al. Tocilizumab therapy reduced intensive care unit admissions and/or mortality in COVID-19 patients. Med Mal Infect. 2020 May 6;S0399-077X(20)30129-3. doi: 10.1016/j.medmal.2020.05.001.

24. Colaneri M, Bogliolo L, Valsecchi P, Sacchi P, Zuccaro V, Brandolino F, et al. Tocilizumab for treatment of severe COVID-19 patients: preliminary results from SMAtteo COvid19 Registry (SMACORE). Microorganisms. 2020 May 9;8(5):E695. doi: 10.3390/microorganisms8050695.

25. Campochiaro C, Delia-Torre E, Cavaili G, De Luca G, Ripa M, Boffini N, et al. Efficacy and safety of tocilizumab in severe COVID-19 patients: a single-center retrospective cohort study. Eur J Intern Med. 2020 May 22; S0953-6205(20)30199-0. doi: 10.1016/j.ejim.2020.05.021.

26. Ruan Q, Yang K, Wang W, Jiang L, Song J. Clinical predictors of mortality due to COVID-19 based on an analysis of data of 150 patients from Wuhan, China. Intensive Care Med. 2020;46(5):846–848.

27. Nishimoto N, Terao K, Mima T, Nakahara H, Takagi N, Kakehi T, et al. Mechanisms and pathologic significances in increase in serum interleukin-6 (IL-6) and soluble IL-6 receptor after administration of an anti-IL-6 receptor antibody, tocilizumab, in patients with rheumatoid arthritis and Castleman disease. Blood 2008;112(10):3959–3964.

28. Slaats J, Ten Oever J, van der Veerdonk FL, Netea MG. IL-1β/IL-6/CRP and IL-18/ferritin: distinct inflammatory programs in infections. PLoS Pathog. 2016;12(12):e1005973

29. Yokota S, Tanaka T, Kishimoto T. Efficacy, safety and tolerability of tocilizumab in patients with systemic juvenile idiopathic arthritis. Ther Adv Musculoskelet Dis. 2012;4(6):387–397.

30. Mazzoni A, Salvati L, Maggi L, Capone M, Vanni A, Spinicci M, et al. Impaired immune cell cytotoxicity in severe COVID-19 is IL-6 dependent. J Clin Invest. 2020 May 28;138554. doi: 10.1172/JCI138554.

